# Virological and Serological Characterization of SARS-CoV-2 Infections Diagnosed After mRNA BNT162b2 Vaccination

**DOI:** 10.1101/2021.09.21.21263882

**Authors:** Francesca Colavita, Silvia Meschi, Cesare Ernesto Maria Gruber, Martina Rueca, Francesco Vairo, Giulia Matusali, Daniele Lapa, Emanuela Giombini, Gabriella De Carli, Martina Spaziante, Francesco Messina, Giulia Bonfiglio, Fabrizio Carletti, Eleonora Lalle, Lavinia Fabeni, Giulia Berno, Vincenzo Puro, Antonino Di Caro, Barbara Bartolini, Giuseppe Ippolito, Maria Rosaria Capobianchi, Concetta Castilletti, on behalf of INMI Covid-19 laboratory surveillance team

## Abstract

Coronavirus disease 2019 (COVID-19) vaccines are proving to be very effective in preventing severe illness; however, although rare, post-vaccine infections have been reported. The present study describes 94 infections (47.9% symptomatic, 52.1% asymptomatic), occurred in Lazio Region (Central Italy) in the first trimester 2021, after first or second dose of mRNA BNT162b2 vaccine. Median viral load at diagnosis was independent from number and time of vaccine dose administration, despite the higher proportion of samples with low viral load observed in fully vaccinated individuals. More importantly, infectious virus was cultured from NPS collected from both asymptomatic and symptomatic vaccinated individuals, suggesting that, at least in principle, they can transmit the infection to susceptible people. The majority of the post-vaccine infections here reported, showed pauci/asymptomatic clinical course, confirming the impact of vaccination on COVID-19 disease. Most cases (78%) showed infection in presence of neutralizing antibodies at the time of infection diagnosis, presumably attributable to vaccination, due to the concomitant absence of anti-N IgG in most cases. The proportion of post-vaccine infections attributed either to Alpha and Gamma VOCs was similar to the proportion observed in the contemporary unvaccinated population in Lazio region. In addition, mutational analysis did not suggest enrichment of a defined set of Spike protein substitutions depending on the vaccination status. Characterization of host and virus factors associated with vaccine breakthrough, coupled with intensive and continuous monitoring of involved viral strains, is crucial to adopt informed vaccination strategies.

## Introduction

In less than 12 months after the beginning of the COVID-19 pandemic, scientific research succeeded in developing multiple vaccines against a previously unknown viral pathogen, severe acute respiratory coronavirus 2 (SARS-CoV-2). The mRNA-based Pfizer-BioNTech vaccine (BNT162b2) has been the first authorized, and on 27 December 2020, the European Union countries launched a co-ordinated vaccination campaign that initially was prioritized for individuals at high risk of SARS-CoV-2 exposure, such as the health-care workers (HCW), and for those at high risk of severe COVID-19, including elderly and residents of assisted living facilities. One key point is to identify and follow up post-vaccine infections and to assess vaccine effectiveness in reducing disease severity and limiting infection transmission to unvaccinated individuals. As expected due to the fact that COVID-19 vaccines do not offer 100% protection against the SARS-CoV-2 infection, current evidence showed that individuals who received full vaccination can become infected ^1^; however, they seem to have lower risk to develop severe COVID-19 compared to unvaccinated people, and to carry reduced viral loads with a likely lower transmission to others ^2–5^. Nevertheless, definitive evidences are still lacking. An additional concern for vaccine effectiveness is about SARS-CoV-2 variants in the Spike protein, which are emerging and spreading across the globe; in fact, some showed potential of immunological escape from the antibodies response that could impact the vaccination efficacy and lead to COVID-19 epidemic rebounds ^6–8^. A specific surveillance on SARS-CoV-2 variants has been established in Lazio Region (Central Italy) in line with the national and international guidelines ^1^. SARS-CoV-2 positivity after vaccination is one of the priority recommendation for genomic characterization in order to identify genetic changes that may be associated with vaccine escape. Here, we described virological and serological testing performed at the Regional Reference Laboratory (RRL) of Virology of the National Institute for Infectious Diseases “L. Spallanzani” (INMI) in Rome, Italy, on samples collected at the time of first RT-PCR positivity from 94 individuals, who resulted positive for SARS-CoV-2 between December 27 and March 30, 2021, following one or two doses of BNT162b2 vaccine.

## Materials and Methods

### Study group

In the frame of the Regional Surveillance programme, naso-pharyngeal swabs (NPS) and, possibly, sera collected from individuals who resulted positive for SARS-CoV-2 after vaccination were sent to INMI in Rome, Italy, for further laboratory investigation. These individuals were tested at peripheral laboratories either following symptoms onset, for contact tracing or screening activities. In this study, we included the first batch of samples (94 NPS) which were referred between December 27 and March 30, 2021 to the RRL at INMI for virological evaluation of post-vaccination RT-PCR positivity occurred at least one day after one or two BNT162b2 vaccine doses and for which in-depth analysis was completed. For 79 individuals, a known date of vaccination was reported and the time lapsing from vaccination to sample collection was calculated. We classified the individuals with known vaccination date into three groups based on the time elapsed from the first dose of vaccine to infection, i.e. time of SARS-CoV-2 positive test or symptoms onset (here considered both as infection starting date): Group 1, individuals tested positive 1 to 15 days after first dose; Group 2, 16 to 30 days after first dose vaccination, and Group 3, >30 days from first dose, 10 days after the second dose injection (considered as full vaccinated group). For 50 individuals (44 of them with reported date of vaccination), a serum sample collected at time of diagnosis was also available for serological testing. In fact, according to the local surveillance system, serum collection was recommended but not mandatory for post-vaccine infections follow-up. NPS and serum samples were shipped to INMI under controlled temperature (−80°C and refrigerated at +4°C, respectively).

Sequencing data (n=1072) produced at INMI exclusively from randomly selected samples collected from unvaccinated individuals during the same study period and representing all the regional territory, were used to evaluate the prevalence of variants of concern (VOCs).

### Molecular testing and virus characterization

Semi-quantitative estimation of viral load was assessed by RT-PCR using DiaSorin Simplexa® COVID-19 Direct platform (DiaSorin, Saluggia, Italy), based on cycle threshold (Ct) value. For whole-genome sequencing, nucleic acid extraction was performed using QiaSymphony automatic extractor and the DPS Virus/Pathogen Midi Kit (QIAGEN), followed by Next-Generation Sequencing (NGS) carried out on Ion Torrent or Illumina Platform using Ion AmpliSeq SARS-CoV-2 Research Panel, following manufacturer’s instructions (ThermoFisher, US). Sequencing was performed on Ion Gene Studio S5 Sequencer and on Illumina MiSeq. Complete genome sequences were obtained combining in-house pipeline ESCA ^9^ with IRMA ^10^ and DRAGEN RNA Pathogen Detection 3.5.15 (Illumina BaseSpace) software, and submitted on GISAID platform ^11^. In case of low coverage for the full-genome characterization, Sanger sequencing was used to fill the NGS gaps in the Spike coding gene.

### Virus isolation

Viral culture was performed in a biosafety level 3 (BSL-3) laboratory at INMI on Vero E6/TMPRSS2 (kindly provided by Dr. Oeda S., National Institute of Infectious Diseases, Tokyo, Japan), as previously described ^12^.

### Serological testing

Serological investigation included the detection of anti-N and anti-RBD Spike IgG using Abbott SARS-CoV-2 assay on Abbott ARCHITECT® i2000sr (Abbott Diagnostics, Chicago, IL, USA) and the evaluation of the neutralising antibodies (nAb) titres using SARS-CoV-2 microneutralization test (MNT) based on live virus ^13^. The viral strain used in MNT (lineage B.1 - clade G; GISAID accession number: EPI_ISL_568579; EVAG Ref-SKU: 008V-04005) had been isolated in March 2020 in Italy and kindly provided by Prof. Baldanti F, Fondazione IRCCS Policlinico San Matteo, Pavia, Italy. SARS-CoV-2 neutralization titers are expressed as the reciprocal of serum dilution achieving 90% reduction of cytopathic effect. First international standard for anti-SARS-CoV-2 provided by the National Institute for Biological Standards and Control (NIBSC, UK) was used as reference in MNT (Research reagent for anti-SARS-CoV-2 human immunoglobulin, NIBSC code: 20/136).

### Statistics

Epidemiological and demographic data were extracted from the Regional Surveillance Information System established by the regional health authority and analyzed using the STATA 14 software. Demographic characteristics of the vaccinated individuals were described using median and interquartile range (IQR) for continuous parameters, and absolute and relative (percentage) frequencies for categorical variables. Inferential analysis of association were performed using chi-square or Fisher exact tests for categorical variables, and Mann–Whitney or Kruskal-Wallis tests for continuous parameters. Univariate analysis using Odd Ratio (OR) with 95% interval of confidence (95%CI) are shown. Analyses were performed using GraphPad Prism version 8.00 (GraphPad Software, La Jolla California) and SPSS V.23 for Windows statistical software; p-value<0.05 was considered statistically significant.

### Ethics

This work was performed within the framework of the COVID-19 outbreak response and surveillance program and the laboratory characterization of post-vaccination infections by INMI laboratory as the RRL, is essential part of the Lazio surveillance regional plan. Use of laboratory and epidemiological records for research purpose has been approved by the INMI Ethical Committee (issue n. 214/20-11-2020), and the need of informed consent form was waived. The study has been conducted in respect of current legislation on personal data protection, all data are presented in non-identifiable form.

### Data availability statement

Sequencing data that support the findings of this study have been deposited in GISAID; the list of the accession codes is available from the corresponding author on reasonable request. The other data used and/or analyzed during the current study are available, only for sections non-infringing personal information, from the corresponding author upon request.

## Results

### Cases of SARS-CoV-2 infections after BNT162b2 vaccination

According to the Regional Surveillance Information System of the Lazio regional health authority, from the 27th of December 2020, the start of the vaccination campaign, up to 30th March 2021, 735,616 individuals received one or two doses of BNT162b2 vaccine. Among these, 1,879 (0.3%) tested positive for SARS-CoV-2 positive at least 1 day after vaccination. The majority (79.5%) of these individuals are reported positive before the second dose injection.

In the study period, samples from 121 out of the reported 1,879 infections in vaccinated persons were recovered from the laboratories that performed the diagnosis, and sent to the RRL to perform the characterization foreseen in the framework of the COVID-19 outbreak response and surveillance program established in Lazio Region. The results described in this study represent those obtained on the first 94 NPS for which analysis was completed. The median time between infection recognition (as symptoms onset or time of first diagnosis for asymptomatic patients) and testing was 0.5 days (range: 0-9 days, with 7 tested samples collected >4 days after diagnosis or symptoms onset).

Demographic and epidemiologic data, including clinical information, are shown in **Table 1**. The majority (n=82, 87.2%) of the 94 individuals under investigation was HCW, the remaining samples were from elderly people (over 80 years old). Median age was 50.5 years old (IQR: 62.0-39.8), 56 (59.6%) were female. Forty-nine (52.1%) were asymptomatic at the diagnosis, and underwent SARS-CoV-2 testing for periodic screening or as contacts of positive cases. The majority (n=61, 64.9%) of post-vaccination cases had a pauci/a-symptomatic clinical course, while a mild disease was reported for 26 (27.7%), and severe for 7 (7.4%) patients. All patients with severe disease had one or more pre-existing co-morbidities, including cardiovascular chronic diseases, diabetes, obesity, renal affections, and neurological disorders, 4 were over 80 years old. Age and co-morbidities were significantly associated with severe disease (p=0.008 and <0.001, respectively). According to the information available at the time of writing (n=85), most infected persons (97.6%) cleared the virus and recovered, while 2 patients died; both dead patients were over 85 years old, presented pre-existing co-morbidities (i.e., cardiovascular chronic diseases, diabetes and neurological disorders), and tested positive after full vaccination (9 and 19 days after the second dose, respectively).

**Table 1.** Demographic and epidemiological information available for the study cohort.

| Characteristic | Total cases<br>tested positive<br>after vaccination<br>(N=94) | Cases with known date of vaccination (N=79) |  |  |  |
| --- | --- | --- | --- | --- | --- |
|  |  | Group 1 | Group 2 | Group 3 | p-value<sup>a</sup> |
|  |  | (n=9,<br>11.4%) | (n=16,<br>20.3%) | (n=54,<br>68.4%) |  |
| Age (median, range) | 55.5 (23-92) | 53 (24-59) | 57.5 (24-82) | 49 (23-87) | 0.540 |
| Gender (n, %) |  |  |  |  |  |
| Male | 38 (40.4%) | 2 (22.2%) | 8 (50.0%) | 22 (40.7%) | 0.397 |
| Female | 56 (59.6%) | 7 (77.8%) | 8 (50.0%) | 32 (59.3%) |  |
| Category (n, %) |  |  |  |  |  |
| HCW | 82 (87.2%) | 9 (100.0%) | 13 (81.3%) | 50 (92.6%) | 0.228 |
| Age over 80 | 12 (12.8%) | - | 3 (18.8%) | 4 (7.4%) |  |
| Symptoms at diagnosis (n, %) |  |  |  |  |  |
| Yes | 45 (47.9%) | 8 (88.9%) | 8 (50.0%) | 23 (42.6%) | 0.037 |
| No | 49 (52.1%) | 1 (11.1%) | 8 (50.0%) | 31 (57.4%) |  |
| Clinical course (n, %) |  |  |  |  |  |
| Pauci/asymptomatic | 61 (64.9%) | 5 (55.5%) | 10 (62.5%) | 38 (70.4%) | 0.208 |
| Mild | 26 (27.7%) | 2 (22.2%) | 6 (37.5%) | 13 (24.1%) |  |
| Severe | 7 (7.4%) | 2 (22.2%) | - | 3 (5.5%) |  |
| Hospitalization (n, %) |  |  |  |  |  |
| Yes | 10 (10.6%) | 2 (22.2%) | 1 (6.3%) | 4 (7.4%) | 0.046 |
| Admission to ICU | 1 (10.0%) | - | - | - |  |
| No | 84 (89.4%) | 7 (77.8%) | 15 (93.7%) | 50 (92.6%) |  |
| COVID-19 outcome (n, %)<sup>#</sup> |  |  |  |  |  |
| Recovered | 83 (97.6%) | 9 (100.0%) | 15 (100.0%) | 46 (95.8%) | 0.598 |
| Dead | 2 (2.4%) | - | - | 2 (4.2%) |  |

<sup>a</sup>By Kruskal-Wallis tests (for quantitative variables) and Chi-squared test (for categorical variables);
p-values <0.05 were considered as statistically significant (in bold).
<sup>#</sup> Available at the time of analysis for: total vaccinees, 85/94; Group 1, 9/9; Group 2, 15/16; Group 3, 48/54.

For 79 individuals, a known date of vaccination was reported **(Table 1)**. The median time between the first-dose vaccination and symptoms onset, or time of first diagnosis for asymptomatic cases, was 47 days, ranging from 1 to 85 days after the first dose (corresponding to 64 days following the full vaccination). Fifty-four (68.4%) individuals resulted infected after full vaccination.

Symptoms at diagnosis and hospitalization rate were significantly more frequent in patients infected after a single dose as compared to patients infected after full vaccination course; a trend towards less frequent severe course was observed in infections acquired after two doses.

### SARS-CoV-2 viral loads and infectivity in NPS from individuals infected up to 2 months after full vaccination

Median Ct values of NPS at diagnosis was 21.2 (IQR: 17.5-31.3), with no significant difference between asymptomatic and symptomatic presentation (median Ct values: 22.0 vs 19.6, **Figure 1A**), even in those patients tested positive after full vaccine course (median Ct values: 21.8 vs 18.9; **Figure 1A**). To understand whether viral RNA was associated to infectiousness, virus isolation was attempted on 84 NPS; 10 NPS were not tested due to due to viral infectivity inactivation by guanidine isothiocyanate contained in the transport medium used for sample collection. Notably, infectious virus was rescued from 44 (52.4%) NPS, 24 collected from symptomatic individuals, 20 collected from asymptomatic subjects. No significant difference in culture positivity rates were found between these two groups (54.5% in symptomatic vs 50.0% in asymptomatic, p=0.827). Similar results were obtained when considering fully vaccinated patients only (60.9% in symptomatic vs 48.2% in asymptomatic, p=0.567). The proportion of samples with low viral load (Ct values >30) in asymptomatic individuals (32.6%) was higher compared to the symptomatic patients (20.0%), but did not reach statistical significance (p=0.242).

**Figure 1.**
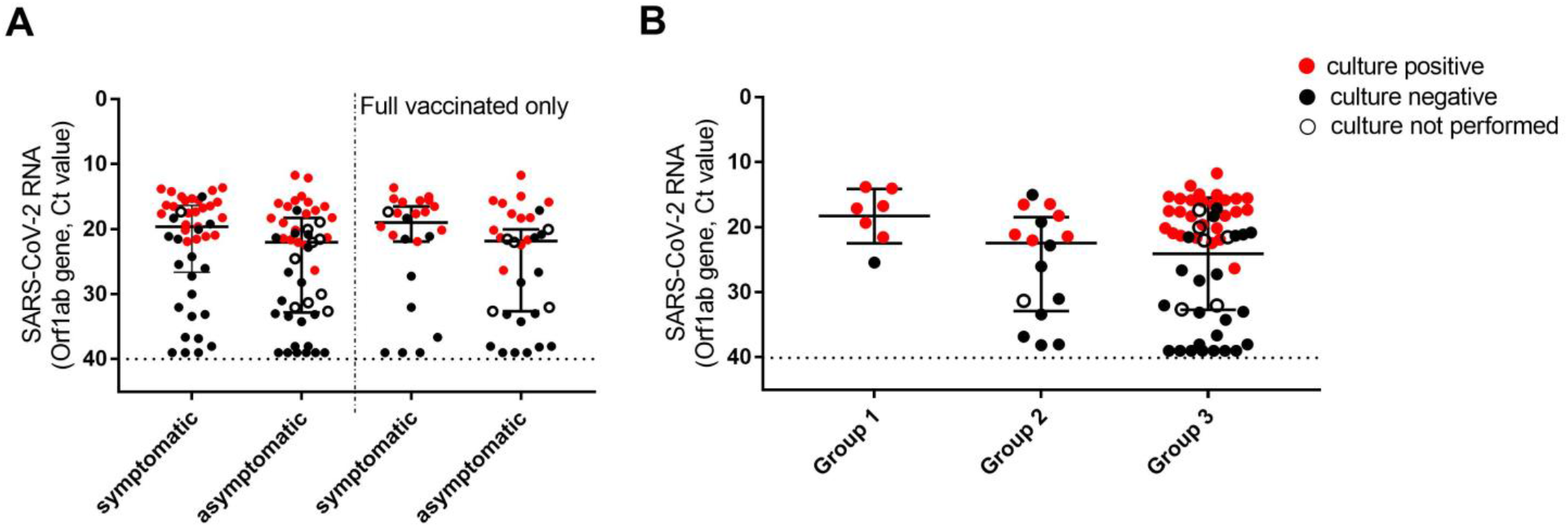
Viral loads and viral culture outcomes in NPS collected in individuals tested positive after first dose vaccination. **A)** Viral loads detected in all symptomatic and asymptomatic individuals at the time of diagnosis (left, n=45 and 49, respectively), in a subgroup including only individuals tested positive after 10 days from second dose vaccination (right; n=24 and 32, respectively). **B)** Viral loads detected in NPS collected at different time points from first dose vaccination. Group 1 (time lapse 1-15 days), n=7; Group 2 (16-30 days), n=16; Group 3 (>30 days), n= 56. Viral RNA levels are expressed as Ct of Orf1ab gene amplification, horizontal dashed line represents the limit of detection of RT-PCR (Ct: 40.0). Median Ct values and IQR are shown. Statistical analysis was performed by Mann–Whitney test, p=0.053 (left) and p=0.098 (right) in (A), and by Kruskal-Wallis test, p=0.135 in (B). Samples yielding positive or negative viral culture are indicated in red and black, respectively; empty dots indicate samples for which viral culture was not performed.

We next considered the viral RNA levels according to time lapsed from first dose to diagnosis, known for 79 individuals, who were divided in 3 groups. The results indicated similar median Ct values in all 3 groups **(Fig. 1B**), despite the higher proportion of samples with low viral load in Group 3. This finding coupled with the isolation rate, which was found higher in samples collected shortly after the vaccination (Group 1), but did not reach statistical difference compared to the other groups (Group 1: 85.7%, Group 2: 40.0%; Group 3: 54.0%, p=0.411). As shown in detail in **Figure 2**, over 39 positive viral cultures, 27 (67.5%) were obtained from fully vaccinated individuals, who tested positive for SARS-CoV-2 up to 85 days after the first vaccination dose. The median Ct value of the samples with positive viral culture was 17.5 (IQR 15.6-20.1), and isolation of infectious virus was strongly associated only with the viral RNA concentration in the NPS, with OR >100 for Ct≤25 vs Ct>25 **(Table 2)**.

**Table 2.**
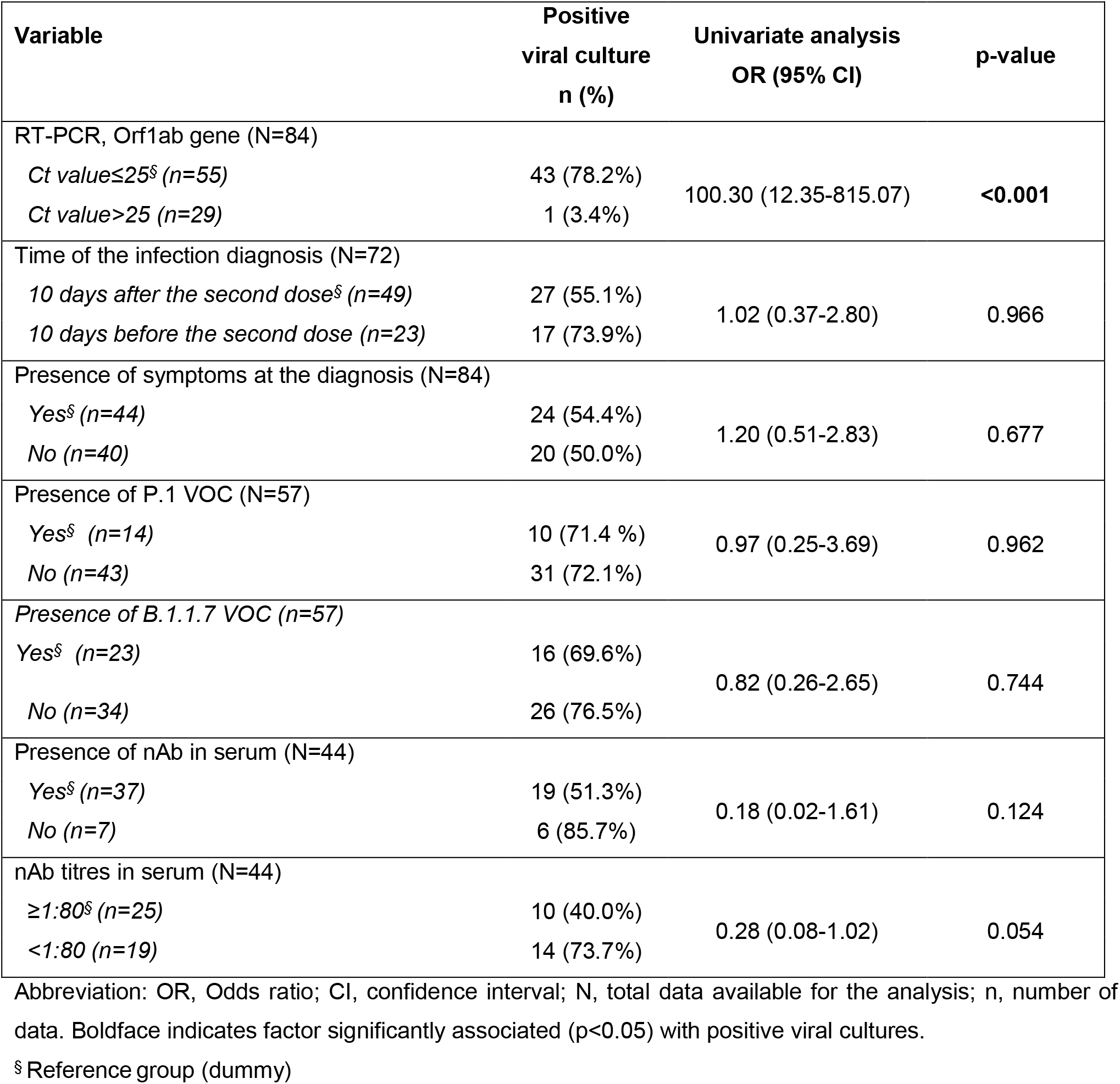
Factors associated with positive viral cultures on NPS collected from vaccinated individuals at the time of SARS-CoV-2 diagnosis.

**Figure 2.**
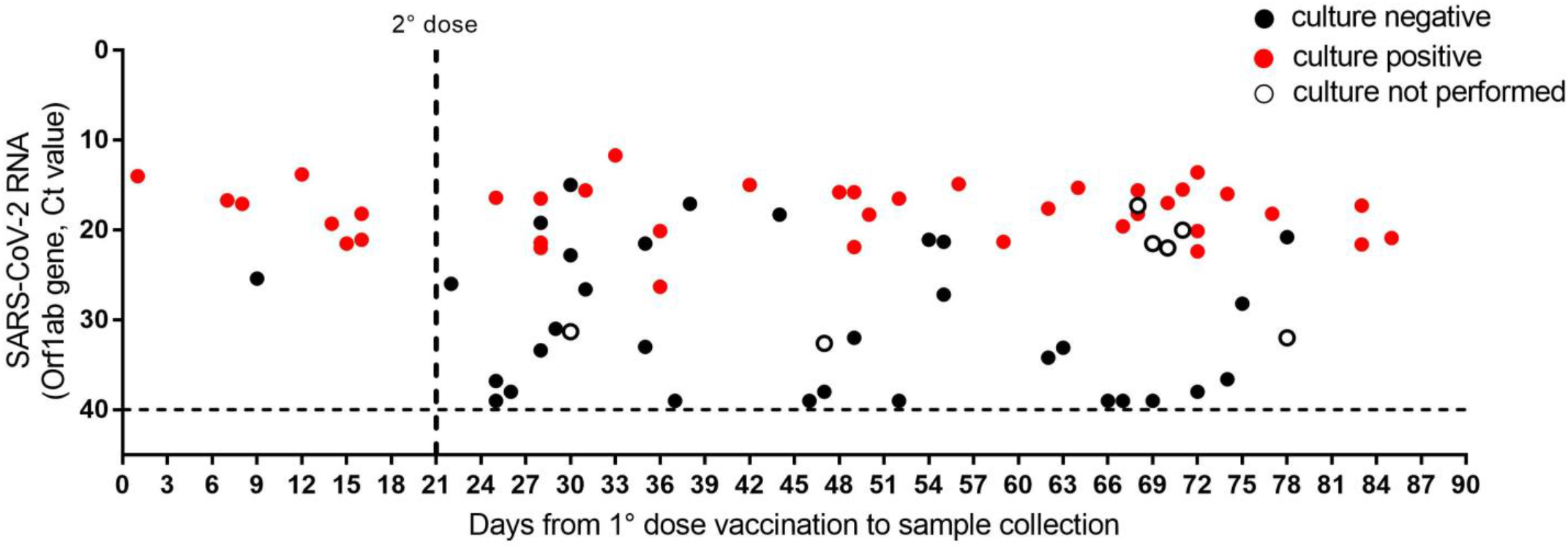
Viral loads and viral culture outcomes in NPS collected in individuals tested positive at different time from the first dose vaccination. Viral RNA levels are expressed as Ct of Orf1ab gene amplification, horizontal dashed lines represent the limit of detection of RT-PCR (Ct: 40). Samples yielding positive or negative viral culture are indicated in red and black, respectively; empty dots indicate samples for which viral culture was not performed. Vertical dashed line represents the time of second dose vaccine administration.

### Antibody response at the time of infection diagnosis

Serological testing was performed on the available serum samples (n=50) which have been collected at the time of diagnosis at the peripheral laboratories and sent to the RRL **(Supplementary Table 1)**. The results showed that antibody response was detected in 48 individuals (96.0%, median anti-RBD Spike IgG BAU/ml=704.6, IQR 403.0-2111.0) at the time of infection diagnosis since first dose vaccination; 42 (84.0%) of them presented also detectable nAb (median titre=1:80, IQR 1:40-1:160), mostly (66.7%) fully vaccinated. Eight vaccinees (16.0%) did not show detectable nAb at diagnosis, of whom 2 (4.0%) were fully vaccinated (both over 80 years old and presenting co-morbidities), indicating primary non response to vaccine. Out of 44 samples collected from individuals with known date of vaccination, we observed significant differences in antibody titres, both anti-RBD Spike IgG and nAb **(Figure 3A-B)**, along the different time-points from vaccination, with high titres after 15 days from the first dose (Groups 2 and 3). Unexpectedly, Group 2 presented higher antibody levels compared to Group 3, which included fully vaccinated individuals. This difference resulted statistically significant for anti-S IgG (p<0.01), not for neutralizing antibodies.

**Figure 3.**
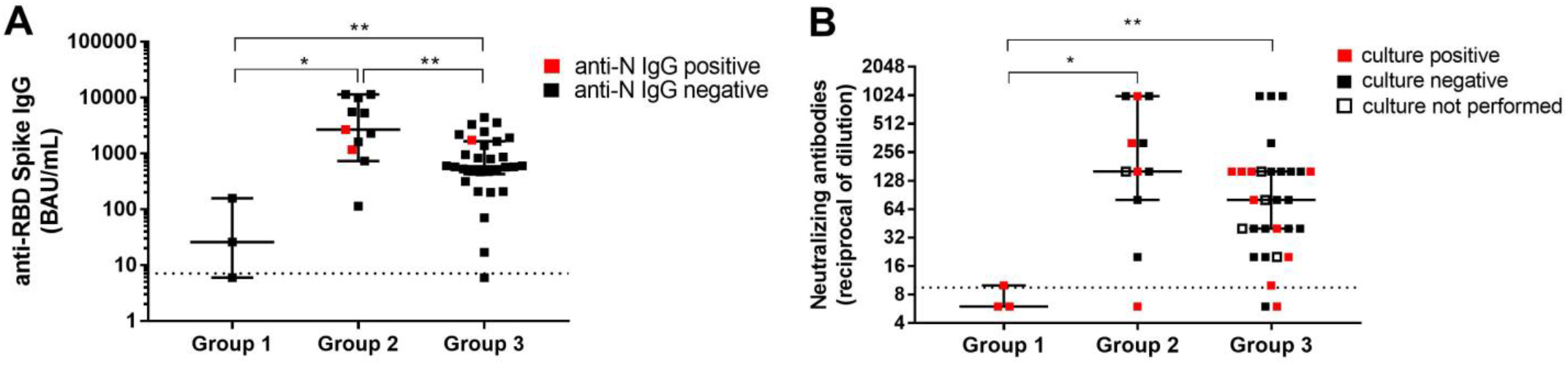
Antibody levels in vaccinated individuals tested positive for SARS-CoV-2. **A)** Antibody titers anti-RBD Spike IgG and **B)** nAb according to the different time-points from first dose vaccination to testing (n=44; Group 1, n=3; Group 2, n=11; Group 3, n= 30). Antibody levels are expressed as BAU/ml in (A) and reciprocal of serum dilution shown on a log2 scale in (B). Dashed line represents the limits of detection of the serological assays (7 BAU/ml in (A) and 1:10 in (B). Median values and IQR are shown, and statistical analysis was performed by Kruskal-Wallis tests, p=0.0007 in (A) and p=0.0089 in (B). Mann–Whitney test for single comparisons was also performed, p values are shown in the graph: *p<0.05; **p<0.01. Samples positive or negative for anti-N IgG are indicated in red and black, respectively. Samples yielding positive or negative viral culture are indicated in red and black, respectively; empty dots indicate samples for which viral culture was not performed.

Since anti-N IgG seroconversion at diagnosis was observed only in 3 patients (6.0%), we reasonably argue that the detected functional antibody response was mostly (39/50, 78.0%) elicited by vaccination, and not by the infection per-se. In addition, we have formal evidence of pre-infection (38 and 60 days before infection) vaccine-induced antibodies response in two patients, with nAb titers of 1:160 at day 15 from full immunization for both infections. As shown in **Figure 3B** and **Supplementary Table 1**, the presence of nAb in serum did not preclude virus isolation from NPS, and titres were not correlated with viral load (median Ct values: 21.5, IQR 16.5-32.2; Spearman r=0.220, p-value=0.124). Notably, data from **Table 2** show a trend to negative association of viral isolation rate with high nAb titers (OR=0.28 for nAb ≥1:80 vs <1:80). Considering the immune status in relation to the clinical course of post-vaccination infections, no statistically significant differences in both anti-RBD Spike IgG and nAb median values were found amongst the different groups of disease severity **(Supplementary Figure 1)**, with no significant association between clinical course and presence or high titres (≥1:80) of nAb (p=0.082 and 0.224, respectively).

### Distribution of VOCs in post-vaccination infections

We next investigated the viral variants infecting the vaccinated individuals included in this study. Whole-genome sequences (WGS) were obtained from the 58 NPS samples with adequately high viral load (Ct<28); additional variant strain identification was obtained for 6 patients by partial Sanger sequencing of the S region. Out of these 63 SARS-CoV-2 sequences, 15 (23.8%) belonged to B.1.177 lineage (GV clade), 14 (22.2%) were Gamma variants (P.1 lineage, GR clade) and 28 (44.4 %) were Alpha variants (B.1.1.7 lineage, GRY clade). One (1.6%) resulted to be Variant of Interest (VOI) Eta (B.1.525 lineage, G clade), and 5 (7.9%) sequences belonged to other clades with 3 sequences to the clade GR (i.e., 2 of the B.1.1 lineage, 1 of the B.1.1.39 lineage) and 2 to the clade G (i.e., 1 of the B.1 lineage and 1 of the B.1.258.17 lineage) **(Supplementary Figure 2A)**. The distribution of variants amongst the vaccinated individuals grouped according to vaccination status (i.e., days from vaccination) was related to the time of infection diagnosis and reflected mostly the circulation of the variants in the general population at the time of the infection **(Supplementary Figure 2B)**. For instance, B.1.1.7 was detected mainly in those vaccinated individuals who tested positive in March 2021, regardless the vaccination status; accordingly, Group 3 showed the predominance of B.1.1.7 as this group included mainly vaccinated individuals who tested positive in March 2021. As matter of fact, when we compared our study population to unvaccinated contemporary population, based on 1072 samples randomly collected from Lazio patients between January and March 2021 and sequenced by NGS and Sanger for surveillance purpose, we observed that variants prevalence in vaccinated individuals followed the circulation in the general population. In fact, as shown in Figure 4, we observed a similar temporal distribution between the two groups, with a clear increase of B.1.1.7 followed by P.1 in both groups, without significant difference for both P.1 (p=0.08 in Chi-square test) and B.1.1.7 lineages (p=0.20 in Chi-square test) **(Figure 4)**. Mutational analysis of Spike protein sequences obtained by NGS showed that signature mutations for the VOCs are observed in all groups, and other changes are found in a minority of patients **(Supplementary Figure 3)**; especially for P.1, none of these minor changes seems to be enriched in Group 3, suggesting no association with more resistant forms.

**Figure 4.**
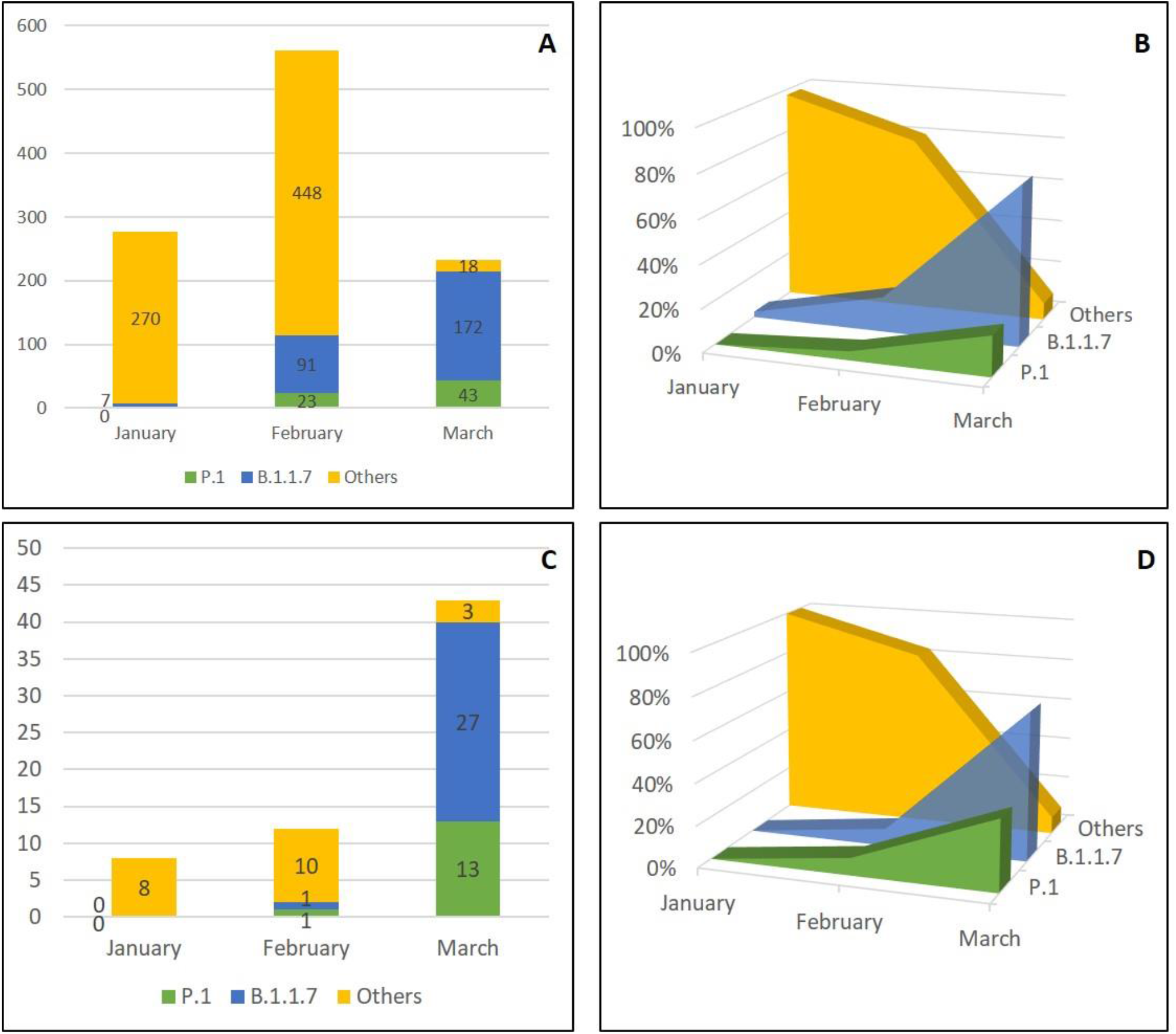
Temporal distribution of PANGO lineages of SARS-CoV-2 genomes. Sequences obtained from unvaccinated individuals (**A-B)** and from vaccinated subjects (**C-D)** in Lazio Region (Central Italy) between January and March 2021 are shown as absolute frequencies (A and C) and percentages (B and D). Others included strains belonging to B.1.177 lineage, B.1.525 lineage, B.1.1 lineage, B.1.1.39 lineage, B.1 lineage, and B.1.258.17 lineage.

The rate of infectious virus isolation in the vaccinated patients was similar when dividing the vaccinated patients according to the infecting virus variants (culture positivity rate: B.1.177; 80.0%; P.1: 71.4%; B.1.1.7, 72.7%; others, 66.7%), with no significant differences in median Ct values amongst the variants (p=0.109) **(Supplementary Figure 4)**. Notably, these findings need to be carefully interpreted since only NSP with high viral loads have been sequenced in order to have an adequate coverage for the full-genome characterization and this could actually bias the analysis.

## Discussion

The present study analyses 94 infections, occurring in Lazio Region (Central Italy) after first or second dose administration of mRNA BNT162b2 vaccine, both from the host side (patients’ demographics, infection severity, antibody status) and from the virus side (viral load, infectivity and infecting variants).

Encouraging evidence is accumulating worldwide about vaccine effectiveness against COVID-19 in real-life vaccination campaigns^14^. Several case-control and population-level studies reported that vaccination substantially reduce both asymptomatic and symptomatic infections, with more significant impact after at least 7 days from the second dose ^2,15–18^. SARS-CoV-2 infections in previously vaccinated individuals are sporadic events compared to the total number of vaccine administrations. Several recent studies including trials and observational reports, showed a lower risk of documented cases in household members of vaccinated HCW and reduced viral load in newly-infected individuals with previous vaccination ^2,5,19^. This has been translated in hypothetical lower infectivity and transmission risk. However, scientific evidence of COVID-19 vaccine effectiveness against infectiousness of SARS-CoV-2 is still limited.

Our data show that vaccinated individuals who get infected after vaccination, although representing a tiny proportion of the vaccinated population (0.3% in our setting), may carry high viral loads in the upper respiratory tract, even when infected long time after the second dose, i.e. when vaccine-related immunity should have been developed. More importantly, we evidenced for the first time that infectious virus can be cultured from NPS collected from both asymptomatic and symptomatic vaccinated individuals, suggesting that they could be able to transmit the infection to susceptible people and potentially be part of transmission chains. As observed for infections in unvaccinated people, high viral load (Ct≤25) is a strong determinant of isolation success ^20,21^. This finding should be carefully considered for public health policy, emphasizing the importance of a proper communication that vaccine does not confer sterilizing immunity; therefore, continued adherence to public health prevention measures is still recommended for vaccinated individuals until adequate vaccination coverage of the population is reached or in presence of susceptible vulnerable individuals ^5,22,23^.On the other hand, the very small proportion of post-vaccine infections is encouraging that overall risk of transmission from vaccinated persons will be low, supporting the crucial role of vaccination.

The impact of emerging variants on the vaccination campaigns is under investigation as vaccine escape variants may be associated to increased vaccine failure ^6^. In the vaccinated individuals described in this study, the majority of infections were caused by Alpha VOC (B.1.1.7), followed by the previously predominant strain in Lazio Region, B.1.177, and by the Gamma VOC (P.1). As recently reported in the United States ^22^, the temporal distribution of the variants identified in the vaccinated individuals clearly matches the pattern of strains circulation in the unvaccinated population of the Lazio Region, with no evidence of vaccine-related immune escape. The preliminary analysis of a more recently collected set of data confirms similar patterns of VOC distribution (both B.1.1.7 and P.1) in vaccinated and unvaccinated individuals from Lazio region, at least up to May 2021, i.e. before the ramp up of the new Delta variant (data not shown).

In a recent study from Israel, Kustin et al. ^24^ reported that infection with Beta VOC was disproportionally detected in fully vaccinated individuals, while Alpha VOC was disproportionally involved in infections diagnosed between 2 weeks after the first dose and 6 days after the second dose. The differences with our results may be due to different pattern of VOCs circulation, as, for example, circulation of Beta VOC was very tiny in our territory. In addition, Spike mutations in each lineage, broken down by Groups 1, 2 and 3, indicated that there is no selection for enrichment of any particular mutation in fully vaccinated individuals (Group 3) as compared to individuals with incomplete vaccination (Groups 1 and 2).

Serological findings showed that the majority (84.0%) of the individuals who tested positive after vaccination had nAb at the time of diagnosis; 64.3% of them, infected after second dose administration, showing relevant nAb titres (≥1:80). Although antibody immune status following vaccination and prior infection was not available for most of these individuals (except for two cases, who underwent serological testing before the infection), in line with recent report ^25^, the nAb titers measured shortly after the infection, together with negative anti-N IgG serology, suggest that it is very likely that they had a immune responses to the vaccine, yet not sufficient to prevent the contagion. On the contrary, 8 individuals tested SARS-CoV-2-positive after vaccination did not present nAb, 2 of them reported as fully vaccinated, suggesting primary non response to vaccine. Unexpectedly, Group 2 presented the highest antibody levels, even compared to Group 3 which included fully vaccinated individuals; while it is reasonable that antibody levels in Group 2 is higher than in Group 1 (due to the longer interval from vaccine administration), the observed difference between Group 2 and 3 seems not to be explained with the available information concerning symptomatic versus asymptomatic infection and distance from symptoms onset, which resulted to be similar between the two groups. A weak response to the vaccination may be a possible explanation. Studies including larger population will help clarify this point. In addition, our data showed that the presence of nAb in serum did not preclude isolation of replication-competent virus from NPS of vaccines; however, the relationship between the infectivity of released virus and the presence of nAb in serum or in the nasopharyngeal tract deserves to be better investigated through dedicated studies.

According to previous reports, the majority of infections observed in vaccinated individuals in this study had a mild or asymptomatic clinical course, and those patients with severe symptoms presented pre-existing co-morbidities and lower median antibody levels compared to those who had a mild or asymptomatic presentation. Two deaths were observed, both in patients over 85 years old and with multiple co-morbidities, which represent a relevant risk factors for severe disease and are prognostic for negative outcome ^4,26–28^. This confirms overall vaccine effectiveness against the disease, which represents a key factor to control morbidity and mortality of SARS-CoV-2 infection, as well as to reduce the public health burden of this pandemic together with social and economic global crisis.

Our study presents some limitation that should be acknowledged. First, the study was an observational study based on real-life data obtained from pandemic surveillance activities, aimed not to establish vaccine efficacy compared to a matched unvaccinated control group, but, to report a virological characterization of those patients reported with SARS-CoV-2 infection despite being vaccinated. Therefore, our observation should be replicated and extended on larger cohorts established *ad hoc* and to other vaccine formulations. Furthermore, no follow-up samples were available for the post-vaccination infected individuals, so that it was not possible to monitor the viral loads and the shedding of infectious virus. In addition, as described above, pre-infection antibody status was only available for few patients. Finally, the evaluation of the cellular immune response would be of great interest to better understand the protection status in cases of vaccine breakthrough infections.

## Acknowledgement

We gratefully acknowledge the Local Health Authorities and the laboratory network (CoroNET) of the Lazio Region. We acknowledge also the contributors of genome sequences of the newly emerging coronavirus (i.e., the originating and submitting laboratories) for sharing their sequences and other metadata through the GISAID Initiative.

## INMI Covid-19 laboratory surveillance team

Abbate Isabella, Agrati Chiara, Agresta Alessandro, Aleo Loredana, Alonzi Tonino, Amendola Alessandra, Apollonio Claudia, Arduini Nicolina, Bartolini Barbara, Berno Giulia, Bettini Aurora, Biancone Silvia, Biava Mirella, Bibbò Angela, Bonfiglio Giulia, Bordi Licia, Brega Carla, Butera Ornella, Canali Marco, Cannas Angela, Capobianchi Maria Rosaria, Carletti Fabrizio, Carrara Stefania, Casetti Rita, Castilletti Concetta, Chiappini Roberta, Ciafrone Lucia, Cimini Eleonora, Coen Sabrina, Colavita Francesca, Condello Rossella, Coppola Antonio, D’Alessandro Gaetano, D’Arezzo Silvia, D’Amato Maurizio, De Santi Claudia, Di Caro Antonino, Di Filippo Stefania, De Giuli Chiara, Fabeni Lavinia, Federici Luigi, Felici Luisa, Ferraioli Valeria, Forbici Federica, Francalancia Massimo, Fusco Maria Concetta, Garbuglia Anna Rosa, Giombini Emanuela, Gramigna Giulia, Graziano Silvia, Gruber Cesare Ernesto Maria, Iazzetti Roberto, Khouri Daniele, Lalle Eleonora, Lapa Daniele, Leone Barbara, Leone Sara, Marafini Giovanni, Marchetti Federica, Massimino Chiara, Matusali Giulia, Mazzarelli Antonio, Meschi Silvia, Messina Francesco, Minosse Claudia, Montaldo Claudia, Mucciante Mirco, Nazzaro Clara, Neri Stefania, Nisii Carla, Petrivelli Elisabetta, Petroni Fabrizio, Petruccioli Elisa, Pisciotta Marina, Pitti Giovanni, Pizzi Daniele, Prota Gianluca, Rozera Gabriella, Rueca Martina, Sabatini Rossella, Santini Francesco, Sarti Silvia, Sberna Giuseppe, Sciamanna Roberta, Scionti Rachele, Selleri Marina, Specchiarello Eliana, Stellitano Chiara, Toffoletti Antonietta, Tonziello Gilda, Truffa Silvia, Turchi Federica, Valli Maria Beatrice, Vantaggio Valentina, Venditti Carolina, Vescovo Tiziana, Vincenti Donatella, Vulcano Antonella, Zambelli Emma.

## Contributions

Study design: C.C., F.C., S.M., B.B., M.R.C., A.D.C.; Laboratory investigation: F.C., S.M., M.R., G.Bo., G.M., D.L., F.C., E.L., L.F., G.Be.; Data collection: F.C., M.R., E.L., G.Be, G.Bo., F.V., M.S., G.D.C., V.P.; Data analysis: F.C., F.V., C.G., E.G., F.M., L.F.; Data interpretation: F.C., C.G., C.C., F.V., M.R.C., A.D.C., Writing: F.C. and M.R.C., with revisions and comments from all authors; Funding acquisition: C.C., A.D.C., M.R.C., G.I.. INMI COVID-19 laboratory surveillance Team contributed to the data and samples collection and storage, and routine diagnostic and epidemiological analyses.

## Supplementary Figures

**Supplementary Figure 1.**
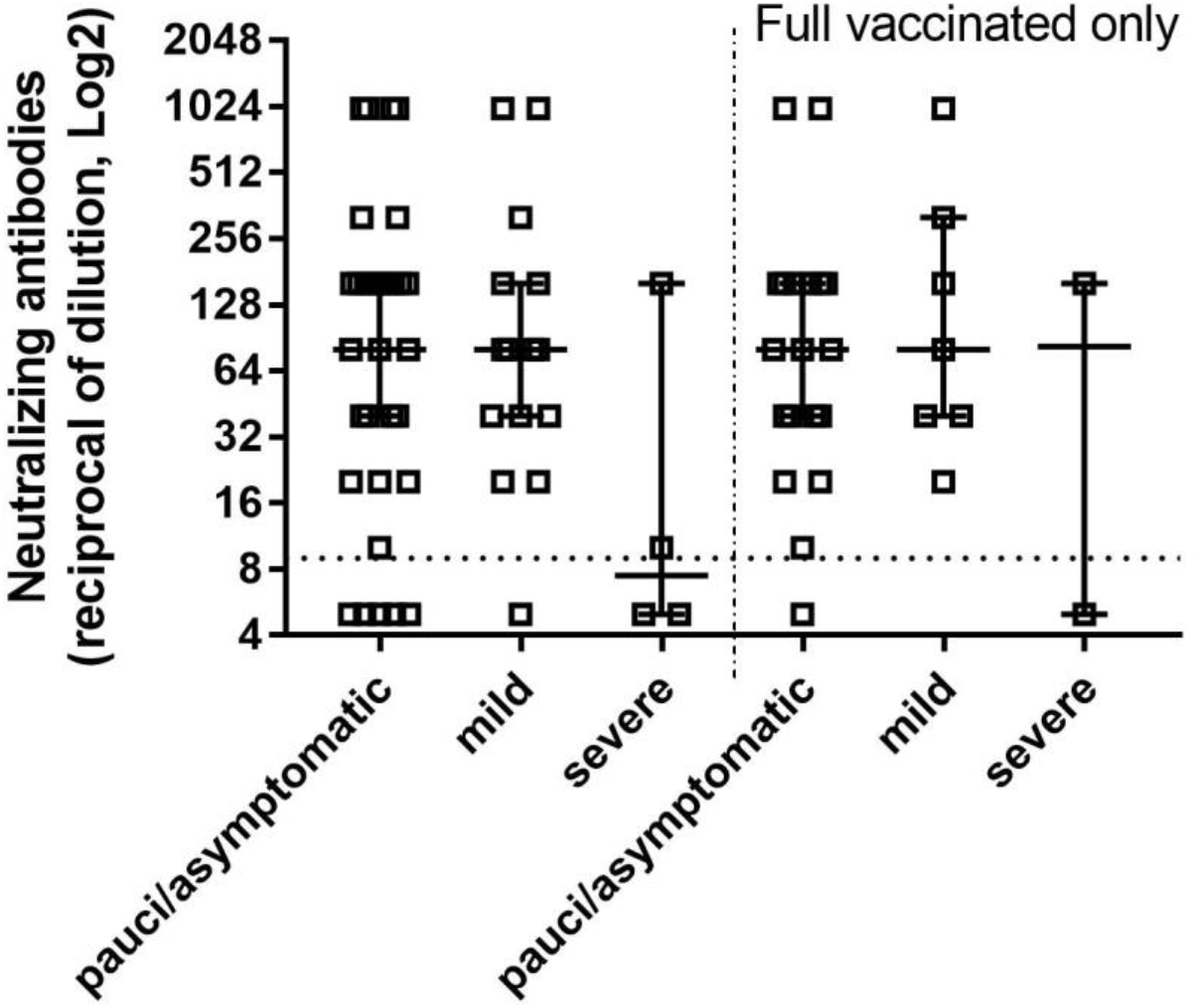
Antibody levels in vaccinated individuals tested positive for SARS-CoV-2 according to the clinical course. Neutralizing antibodies detected in all individuals classified as pauci/asymptomatic (n=31), mild (n=15), and severe (n=4) are shown on the left, those detected only in infections diagnosed after 10 days from second dose vaccination are shown on the right (pauci/asymptomatic, n=19; mild, n=7; severe, n=2). nAb levels are expressed as the reciprocal of serum dilution. Dashed line represents the limits of detection of 1:10. Median values and IQR are shown, and statistical analysis was performed by Kruskal-Wallis test, p=0.220 (left) and p=0.556 (right).

**Supplementary Figure 2.**
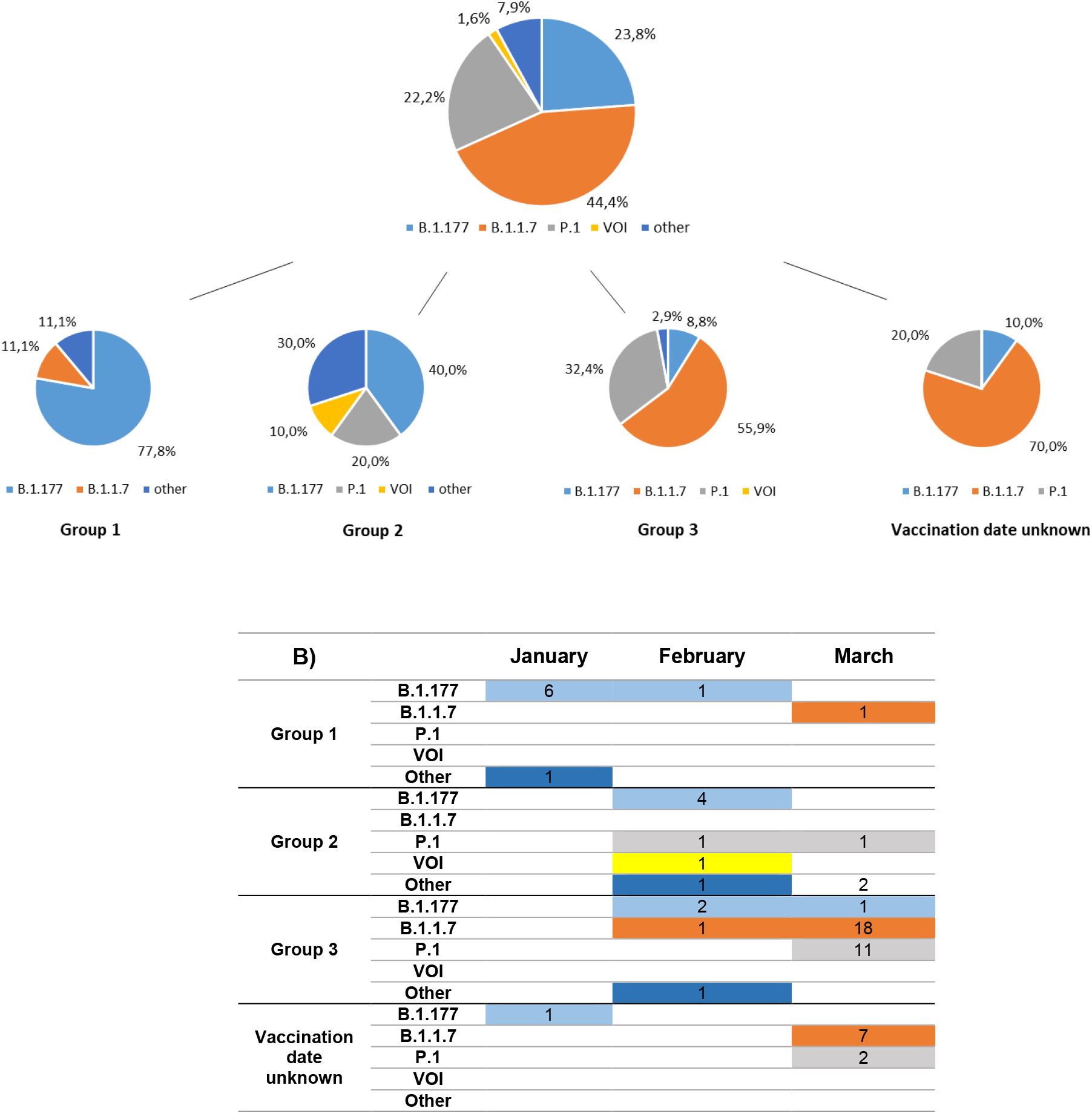
SARS-CoV-2 strains detected in NPS samples collected from post-vaccination infections. **A)** Percentages over a total of 63 sequences obtained are shown. Viral lineages detected in all vaccinated individuals (pie chart above) and divided according to the time elapsed from the first dose of vaccine to infection diagnosis (pie charts below) are indicated: Group 1 (time lapse 1-15 days), n=9; Group 2 (16-30 days), n=10; Group 3 (>30 days), n=34; Unknown, vaccination date not available, n=10. B) Absolute frequencies of viral lineages sequenced from the different vaccinated groups according to the time (month 2021) of infection diagnosis. VOI included B.1.525 lineage; Other includes strains belonging to B.1.1 lineage, B.1.1.39 lineage, B.1 lineage, and B.1.258.17 lineage.

**Supplementary Figure 3.**
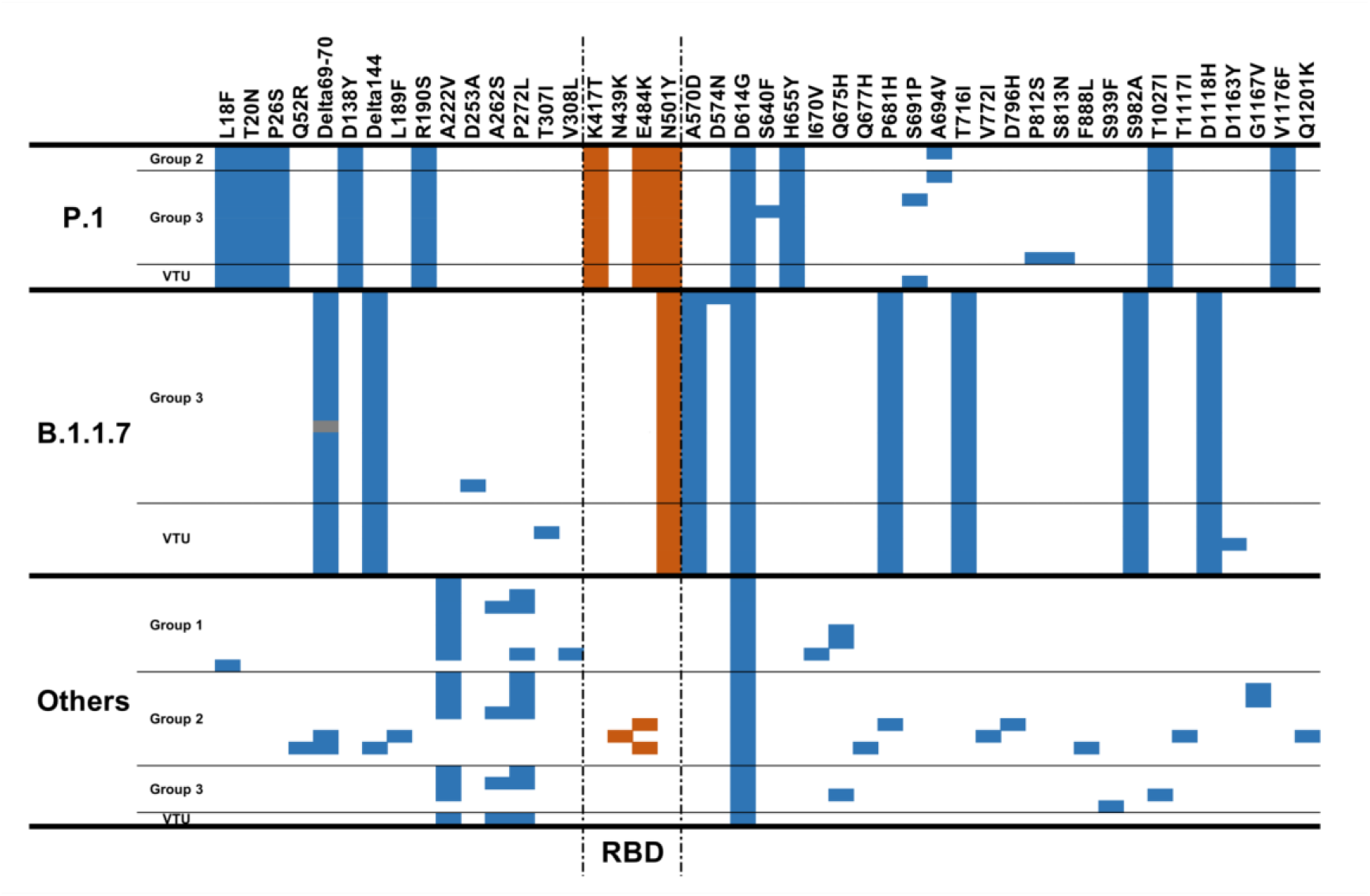
Amino acid substitutions found in the Spike protein of SARS-CoV-2 sequences obtained from NGS analysis of vaccinated individuals. Mutations are shown according to the different vaccinated groups identified on the basis of the time elapsed from the first dose of vaccine to testing: Group 1 (time lapse 1-15 days), n=8; Group 2 (16-30 days), n=10; Group 3 (>30 days), n=30; VTU: Vaccination time unknown, n=9. Mutations found in the receptor-binding domain (RBD) sequence are reported in light-red colour; mutations that cannot be confirmed nor excluded due to low coverage are reported in grey colour.

**Supplementary Figure 4.**
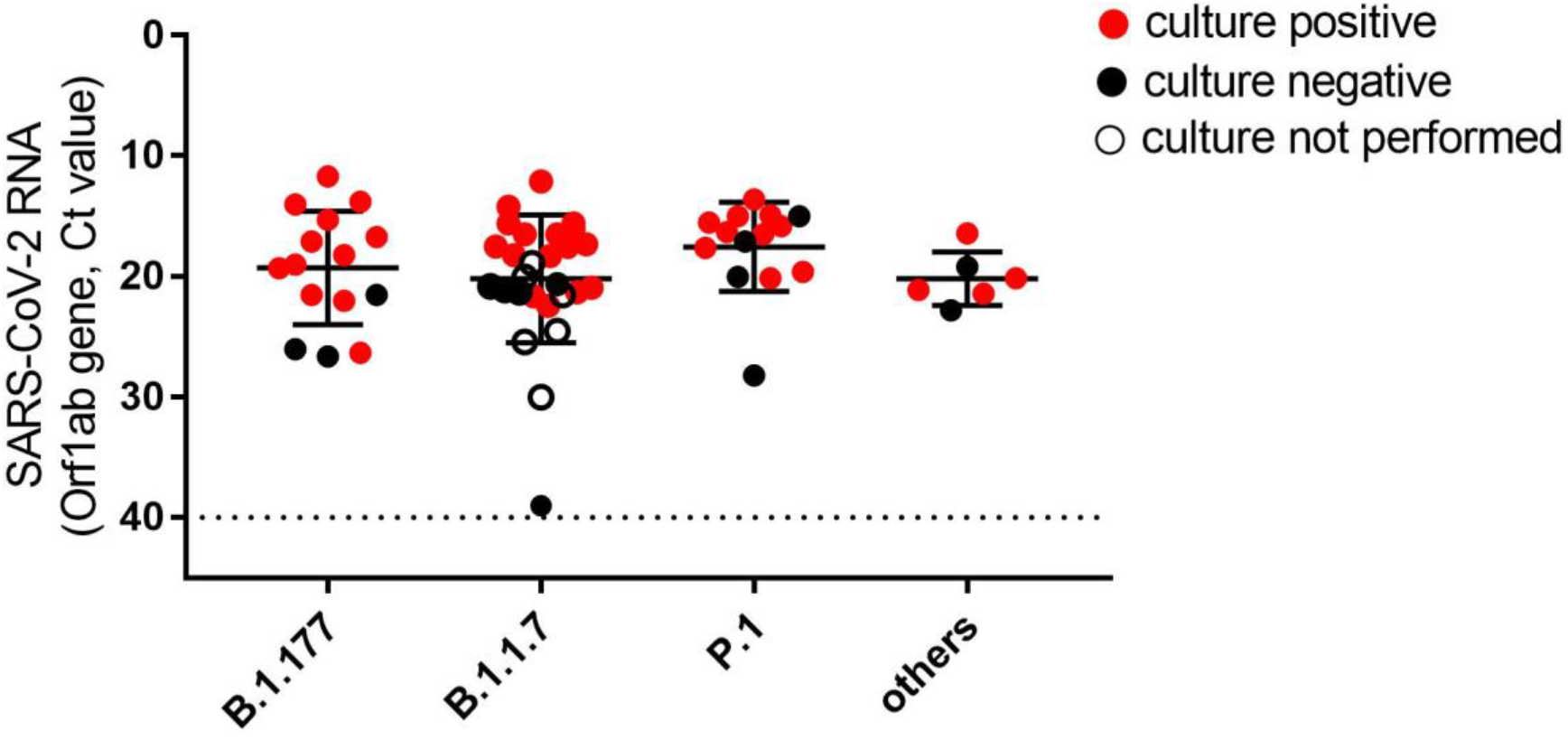
Viral loads and viral culture outcomes in NPS collected in individuals tested positive after first dose vaccination according to the infecting SARS-CoV-2 variants. Viral RNA levels are expressed as cycle threshold values (Ct) of Orf1ab gene amplification, horizontal dashed line represents the limit of detection of RT-PCR (Ct: 40). Median Ct values and IQR are shown (n=63; B.1.177, n=15; B.1.1.7, n=28; P.1, n=14; other, n=6), and statistical analysis was performed by Kruskal-Wallis test, p=0.109. Samples yielding positive or negative viral culture are indicated in red and black, respectively; empty dots indicate samples for which viral culture was not performed.

**Supplementary Table 1.** Vaccinated individuals with available matching data on antibody and RNA viral levels.

| ID | Viral load<br>(Ct values) | Viral<br>culture | Viral<br>Strain | anti-S IgG<br>(BAU/mL<br>≥7.1) | anti-N IgG<br>(Index ≥1,4) | nAb<br>(VNT90≥1:10) | DTV | DSV |
| --- | --- | --- | --- | --- | --- | --- | --- | --- |
| 1 | 17.1 | Positive | B.1.177 | <7.1 | 0.02 | <1:10 | 8 | n.a. |
| 2 | 19.3 | Positive | B.1.177 | 158.2 | 0.02 | <1:10 | 14 | 14 |
| 3 | 21.5 | Positive | B.1.177 | 25.9 | 0.02 | 1:10 | 15 | 12 |
| 4 | 18.2 | Positive | B.1.177 | 1130.6 | 0.02 | <1:10 | 16 | 13 |
| 5 | 26.0 | Negative | B.1.177 | 5549.1 | 0.56 | 1:160 | 22 | 22 |
| 6 | 16.4 | Positive | B.1.525 | 11360.0 | 0.16 | 1:1260 | 25 | 21 |
| 7 | 36.8 | Negative | n.p. | 11360.0 | 1.08 | 1:1260 | 25 | 21 |
| 8 | 38.0 | Negative | n.p. | 1615.2 | 0.11 | 1:80 | 26 | 24 |
| 9 | 33.4 | Negative | n.p. | 733.2 | 0.03 | 1:20 | 28 | 24 |
| 10 | 22.0 | Positive | B.1.177 | 5306.6 | 0.02 | 1:160 | 28 | n.a. |
| 11 | 16.5 | Positive | P.1 | 2.284.60 | 0.56 | 1:320 | 28 | n.a. |
| 12 | 31.0 | Negative | n.p. | 2678.0 | 6.47 | 1:1260 | 29 | n.a. |
| 13 | 31.3 | n.p. | n.p. | 1174.1 | 4.35 | 1:160 | 30 | n.a. |
| 14 | 15.0 | Negative | P.1 | 9889.7 | 0.05 | 1:320 | 30 | 27 |
| 15 | 15.6 | Positive | B.1.1.7 | 17.1 | 0.02 | <1:10 | 31 | 31 |
| 16 | 26.6 | Negative | B.1.177 | 4397.2 | 0.81 | 1:1260 | 31 | n.a. |
| 17 | 11.7 | Positive | B.1.177 | 3311.7 | 0.11 | 1:80 | 33 | n.a. |
| 18 | 26.3 | Positive | B.1.177 | 208.8 | 0.01 | 1:160 | 36 | n.a. |
| 19 | 20.1 | Positive | B.1.1.39 | 1632.8 | 0.03 | 1:160 | 36 | n.a. |
| 20 | 39.0 | Negative | B.1.1.7 | <7.1 | 0.02 | <1:10 | 37 | 37 |
| 21 | 17.1 | Negative | P.1 | 799.1 | 0.19 | 1:80 | 38 | n.a. |
| 22 | 38.0 | Negative | n.p. | 313.7 | 0.17 | 1:1260 | 47 | n.a. |
| 23 | 32.6 | n.p. | n.p. | 2453.0 | 0.07 | 1:80 | 47 | n.a. |
| 24 | 15.8 | Positive | n.p. | 950.8 | 0.01 | 1:80 | 48 | n.a. |
| 25 | 21.9 | Positive | n.p. | 467.3 | 0.05 | 1:40 | 49 | 45 |
| 26 | 15.8 | Positive | P.1 | 861.7 | 0.07 | 1:320 | 49 | 46 |
| 27 | 32.0 | Negative | n.p. | 568.2 | 0.04 | 1:40 | 49 | 47 |
| 28 | 18.3 | Positive | B.1.1.7 | 2179.8 | 0.04 | 1:160 | 50 | n.a. |
| 29 | 27.2 | Negative | n.p. | 825.7 | 0.17 | 1:160 | 55 | 53 |
| 30 | 21.3 | Positive | B.1.1.7 | 575.0 | 0.08 | 1:40 | 59 | n.a. |
| 31 | 15.3 | Positive | B.1.177 | 200.8 | 0.03 | 1:40 | 64 | 57 |
| 32 | 39.0 | Negative | n.p. | 529.8 | 0.01 | 1:20 | 66 | n.a. |
| 33 | 39.0 | Negative | n.p. | 1379.6 | 0.06 | 1:160 | 67 | n.a. |
| 34 | 15.6 | Positive | B.1.1.7 | 70.2 | 0.02 | 1:10 | 68 | n.a. |
| 35 | 17.5 | Positive | B.1.1.7 | 599.1 | 0.05 | 1:160 | 68 | 64 |
| 36 | 39.0 | Negative | n.p. | 1742.9 | 2.55 | 1:160 | 69 | n.a. |
| (continued next page) |  |  |  |  |  |  |  |  |
| 37 | 21.5 | n.p. | B.1.1.7 | 475.0 | 0.04 | 1:20 | 69 | n.a. |
| 38 | 17.3 | Positive | B.1.1.7 | 208.8 | 0.01 | 1:160 | 70 | 67 |
| 39 | 22.0 | n.p. | P.1 | 1903.1 | 0.02 | 1:160 | 70 | n.a. |
| 40 | 13.6 | Positive | P.1 | 574.7 | 0.03 | 1:40 | 72 | 70 |
| 41 | 38.0 | Negative | n.p. | 3569.7 | 0.49 | 1:1260 | 72 | n.a. |
| 42 | 36.6 | Negative | n.p. | 597.9 | 0.1 | 1:20 | 74 | 73 |
| 43 | 32.0 | n.p. | n.p. | 481.2 | 0.54 | 1:40 | 78 | n.a. |
| 44 | 21.6 | Positive | B.1.1.7 | 506.8 | 0.01 | 1:20 | 83 | n.a. |
| 45 | 20.0 | Negative | P.1 | 676.6 | 0.02 | 1:80 | n.a. | n.a. |
| 46 | 18.9 | n.p. | B.1.1.7 | 69.6 | 0.03 | <1:10 | n.a. | n.a. |
| 47 | 16.3 | Positive | P.1 | 732.6 | 0.02 | 1:40 | n.a. | n.a. |
| 48 | 14.2 | Positive | B.1.1.7 | 382.6 | 0.02 | <1:10 | n.a. | n.a. |
| 49 | 33.1 | Negative | n.p. | 609.5 | 0.3 | 1:80 | n.a. | n.a. |
| 50 | 12.1 | Positive | B.1.1.7 | 36.3 | 0.02 | <1:10 | n.a. | n.a. |

Abbreviation: DTV, days from first dose vaccination to testing; DSV, days from first dose vaccination to symptoms onset (for symptomatic individuals only); n.p., not performed; n.a., not available, as vaccination date unknown, or absence of symptoms at diagnosis.

## References

1. European Centre for Disease Prevention and Control. Risk of SARS-CoV-2 transmission from newly-infected individuals with documented previous infection or vaccination. 29th March (2021). Available at: https://www.ecdc.europa.eu/en/publications-data/sars-cov-2-transmission-newly-infected-individuals-previous-infection. (Accessed: 6th May 2021)

2. Levine-Tiefenbrun, M. et al. Initial report of decreased SARS-CoV-2 viral load after inoculation with the BNT162b2 vaccine. Nat. Med. (2021). doi:10.1038/s41591-021-01316-7

3. Kawasuji, H. et al. Transmissibility of COVID-19 depends on the viral load around onset in adult and symptomatic patients. PLoS One 15, e0243597 (2020).

4. Dagan, N. et al. BNT162b2 mRNA Covid-19 Vaccine in a Nationwide Mass Vaccination Setting. N. Engl. J. Med. 384, 1412–1423 (2021).

5. Teran, R. A. et al. Postvaccination SARS-CoV-2 Infections Among Skilled Nursing Facility Residents and Staff Members — Chicago, Illinois, December 2020–March 2021. MMWR. Morb. Mortal. Wkly. Rep. 70, 632–638 (2021).

6. Hoffmann, M. et al. SARS-CoV-2 variants B.1.351 and P.1 escape from neutralizing antibodies. Cell 184, 2384–2393.e12 (2021).

7. Fontanet, A. et al. SARS-CoV-2 variants and ending the COVID-19 pandemic. Lancet (London, England) 397, 952–954 (2021).

8. Wu, K. et al. Serum Neutralizing Activity Elicited by mRNA-1273 Vaccine. N. Engl. J. Med. 384, 1468–1470 (2021).

9. Rueca, M. et al. ESCA pipeline: Easy-to-use SARS-CoV-2 genome Assembler. bioRxiv 2021.05.21.445156 (2021). doi:10.1101/2021.05.21.445156

10. Shepard, S. S. et al. Viral deep sequencing needs an adaptive approach: IRMA, the iterative refinement meta-assembler. BMC Genomics 17, 708 (2016).

11. Elbe, S. & Buckland-Merrett, G. Data, disease and diplomacy: GISAID’s innovative contribution to global health. Glob. challenges (Hoboken, NJ) 1, 33–46 (2017).

12. Colavita, F. et al. Virological Characterization of the First 2 COVID-19 Patients Diagnosed in Italy: Phylogenetic Analysis, Virus Shedding Profile From Different Body Sites, and Antibody Response Kinetics. Open Forum Infect. Dis. 7, (2020).

13. Matusali, G. et al. SARS-CoV-2 Serum Neutralization Assay: A Traditional Tool for a Brand-New Virus. Viruses 13, 655 (2021).

14. Shilo, S., Rossman, H. & Segal, E. Signals of hope: gauging the impact of a rapid national vaccination campaign. Nat. Rev. Immunol. 21, 198–199 (2021).

15. Hall, V. J. et al. Do Antibody Positive Healthcare Workers Have Lower SARS-CoV-2 Infection Rates than Antibody Negative Healthcare Workers? Large Multi-Centre Prospective Cohort Study (The SIREN Study), England: June to November 2020. medRxiv (2021). doi:10.2139/ssrn.3768524

16. Keehner, J. et al. SARS-CoV-2 Infection after Vaccination in Health Care Workers in California. N. Engl. J. Med. 384, 1774–1775 (2021).

17. Thompson, M. G. et al. Interim Estimates of Vaccine Effectiveness of BNT162b2 and mRNA-1273 COVID-19 Vaccines in Preventing SARS-CoV-2 Infection Among Health Care Personnel, First Responders, and Other Essential and Frontline Workers - Eight U.S. Locations, December 2020-March. MMWR. Morb. Mortal. Wkly. Rep. 70, 495–500 (2021).

18. Haas, E. J. et al. Impact and effectiveness of mRNA BNT162b2 vaccine against SARS-CoV-2 infections and COVID-19 cases, hospitalisations, and deaths following a nationwide vaccination campaign in Israel: an observational study using national surveillance data. Lancet (2021). doi:10.1016/S0140-6736(21)00947-8

19. Shah, A. et al. Effect of vaccination on transmission of COVID-19: an observational study in healthcare workers and their households. medRxiv 2021.03.11.21253275 (2021).

20. Wölfel, R. et al. Virological assessment of hospitalized patients with COVID-2019. Nature 581, 465–469 (2020).

21. Jaafar, R. et al. Correlation Between 3790 Quantitative Polymerase Chain Reaction– Positives Samples and Positive Cell Cultures, Including 1941 Severe Acute Respiratory Syndrome Coronavirus 2 Isolates. Clin. Infect. Dis. (2020). doi:10.1093/cid/ciaa1491

22. Birhane, M. et al. COVID-19 Vaccine Breakthrough Infections Reported to CDC — United States, January 1–April 30, 2021. MMWR. Morb. Mortal. Wkly. Rep. 70, 792–793 (2021).

23. Vilches, T. N., Zhang, K., Van Exan, R., Langley, J. M. & Moghadas, S. M. Projecting the impact of a two-dose COVID-19 vaccination campaign in Ontario, Canada. Vaccine 39, 2360–2365 (2021).

24. Kustin, T. et al. Evidence for increased breakthrough rates of SARS-CoV-2 variants of concern in BNT162b2-mRNA-vaccinated individuals. Nat. Med. (2021). doi:10.1038/s41591-021-01413-7

25. Hacisuleyman, E. et al. Vaccine Breakthrough Infections with SARS-CoV-2 Variants. N. Engl. J. Med. NEJMoa2105000 (2021). doi:10.1056/NEJMoa2105000

26. Rydyznski Moderbacher, C. et al. Antigen-Specific Adaptive Immunity to SARS-CoV-2 in Acute COVID-19 and Associations with Age and Disease Severity. Cell 183, 996–1012.e19 (2020).

27. Williamson, E. J. et al. Factors associated with COVID-19-related death using OpenSAFELY. Nature 584, 430–436 (2020).

28. Sanyaolu, A. et al. Comorbidity and its Impact on Patients with COVID-19. SN Compr. Clin. Med. 1–8 (2020). doi:10.1007/s42399-020-00363-4

